# Using aggregate patient data at the bedside via an on-demand consultation service

**DOI:** 10.1101/2021.06.16.21259043

**Authors:** Alison Callahan, Saurabh Gombar, Eli M. Cahan, Kenneth Jung, Ethan Steinberg, Vladimir Polony, Keith Morse, Robert Tibshirani, Trevor Hastie, Robert Harrington, Nigam H. Shah

**Author notes:** Corresponding author: Alison Callahan. Equal contributors (Alison Callahan, Saurabh Gombar).

## Abstract

Using evidence derived from previously collected medical records to guide patient care has been a long standing vision of clinicians and informaticians, and one with the potential to transform medical practice. As a result of advances in technical infrastructure, statistical analysis methods, and the availability of patient data at scale, an implementation of this vision is now possible. Motivated by these advances, and the information needs of clinicians in our academic medical center, we offered an on-demand consultation service to derive evidence from patient data to answer clinician questions and support their bedside decision making. We describe the design and implementation of the service as well as a summary of our experience in responding to the first 100 requests. Consultation results informed individual patient care, resulted in changes to institutional practices, and motivated further clinical research. We make the tools and methods developed to implement the service publicly available to facilitate the broad adoption of such services by health systems and academic medical centers.

## The need for on-demand evidence

Evidence-based medicine emphasizes the “conscientious, explicit and judicious use of current best evidence”^1^ when making treatment decisions^2,3^. Randomized controlled trials (RCTs) are considered the highest quality source of evidence about treatment efficacy and safety. Evidence derived from RCTs, however, often does not generalize to the vast majority of patients, who tend to have multiple comorbidities, take many medications, and differ from individuals enrolled in RCTs on many characteristics^4^, resulting in an inferential gap between the evidence that is available and that which is needed^5,6^. Therefore, it is necessary to transform the evidence generation process^7^ and to incorporate the use of aggregate patient data at the point of care^8^ in order to create a successful learning health system^9^.

Electronic medical records (EMRs) are a source of rich longitudinal data about *millions* of real world patients. Since the 1970s, clinicians and scientists have envisioned using the medical records of previously treated patients to inform the care of current and future patients^10,11^. As a recent example, in 2011 the New England Journal of Medicine published an article by Frankovich et al.^12^ describing the use of EMR data to support management of an adolescent female with systemic lupus erythematosus. At the time, incorporating data from EMRs into clinical decision making required significant manual effort, rendering it infeasible for use in routine patient care.

A decade later, the adoption of EMRs across the United States and internationally, the increasing ease of use of advanced statistical methods, and the ability to compute with large patient cohorts has enabled a core tenet of the learning health system: deriving on-demand evidence for diverse clinical scenarios from the EMR^7,13^.

Using these advances as a foundation, we designed, developed, and offered a consultation service that used EMR and medical insurance claims data at Stanford Medicine to provide on-demand evidence for questions arising during clinical care^14^. Here, we report our findings from responding to the first 100 requests to the service: we summarize requests by medical specialty, the types of analyses required to fulfill their requests, and clinicians’ responses to the evidence returned.

## The setup of the consultation service

Beginning in 2017, with approval from the Stanford Institutional Review Board, we offered a consultation service to provide on-demand evidence to clinicians at Stanford Medicine, staffed by a team of four (described below). As part of offering the service, we collected data on the motivations for consultation requests, and the subsequent actions taken in light of the evidence returned. At the conclusion of the study in August 2019, we analyzed the consultation request motivations and resulting actions, and assessed the concordance of consultation results across clinical data sources as a measure of reliability of consultation analysis methods.

In designing the service, we leveraged best practices^15^, methods^16^, and tools^17,18^ to derive evidence from EMRs. Callahan et al^15^ summarizes recommendations for conducting and reporting observational studies done using EMRs derived from a large body of our team’s prior work. For example, we have used EMR data for *vigilance*, such as monitoring adverse drug events^19,20^ and surveilling implantable devices^21^ ; for *answering clinical questions* such as whether there is an association between androgen deprivation therapy and dementia^22,23^ ; and for *elucidating quality of care*, by profiling unplanned ED visits^24^, surfacing patient reported outcomes^25^ and quantifying treatment variability in metastatic breast cancer^26^. We have also learned from leading collaborative studies^27^, developing methods for electronic phenotyping^28–31^, and from participating in multiple OHDSI network studies^32–38^.

Gombar et al.^14^ describes the consultation service setup to receive questions from clinicians, retrieve the appropriate patient data using a specialized search engine^18^, perform the analyses required for the question, and return a report summarizing the results. Schuler et al.^16^ describes the methods for data extraction, processing, and analysis used in the consultation service. Datta et al^17^ describes the platform for clinical data science at Stanford Medicine that supported the operation of the service.

## The workflow for fulfilling a consultation request

A consultation began with an email from a requestor, detailing a clinical question. Upon receiving the request, the team’s informatics clinician scheduled an intake discussion with the requesting clinician to specify the <u>p</u>opulation, intervention, <u>c</u>omparator, <u>o</u>utcome and timeframe (PICOT) for their particular question^14^.

Based on the PICOT formulation of the question, the EMR data specialist constructed patient cohorts using the Advanced Cohort Engine (ACE)^18^ to search one of three data sources: EMRs of 3.1 million individuals from Stanford Medicine; IBM MarketScan® insurance claims for 124 million individuals; or Optum Clinformatics Data Mart® insurance claims for 53 million individuals. The data scientist then conducted the necessary statistical analyses and worked with the informatics clinician to write a report summarizing the analyses and their results. The report was then shared with the requestor and explained during an in-person debrief session. Each report consisted of the original question as posed, the PICOT re-formulation, and sections summarizing the cohort demographics, the interpretation of the analyses, and a detailed walkthrough of the analyses. Three example reports are provided in the Supplement. The interaction was designed to be similar to obtaining a second opinion from a colleague.

Our workflow evolved to incorporate real-time searches of the EMR as the informatics clinician collected PICOT details. For example, if a given cohort criterion returned very few patients, then the informatics clinician could relay this information during the intake interview in order to elicit modifications to the cohort definition from the requestor. Clarifications needed during debrief interviews were also incorporated into subsequent reports and debriefs to better contextualize analysis results for requestors. The majority of this evolution occurred during the first 3 months of offering the service.

Based on the time required to respond to the first 100 consultations received (see *Findings from the first 100 consultations*), we believe a team composed of one full-time clinical informatics fellow, two full-time EMR data specialists, and a 20% part-time data scientist would be able to complete up to 20 such consultations in one week. The personnel costs for our geographic area (San Francisco Bay) for this team are estimated at $505,000/year. Yearly data access infrastructure, cloud compute, licensing, and professional service expenses come to an additional $70,000/year. With these assumptions, the cost of running such a service would be approximately $550 per consultation.

Figure 1 illustrates the process of fulfilling a consultation request. The datasets, cohort building and analysis methods used in completing consultations, and our assessments of their performance, are further described in the following subsections.

**Figure 1.**
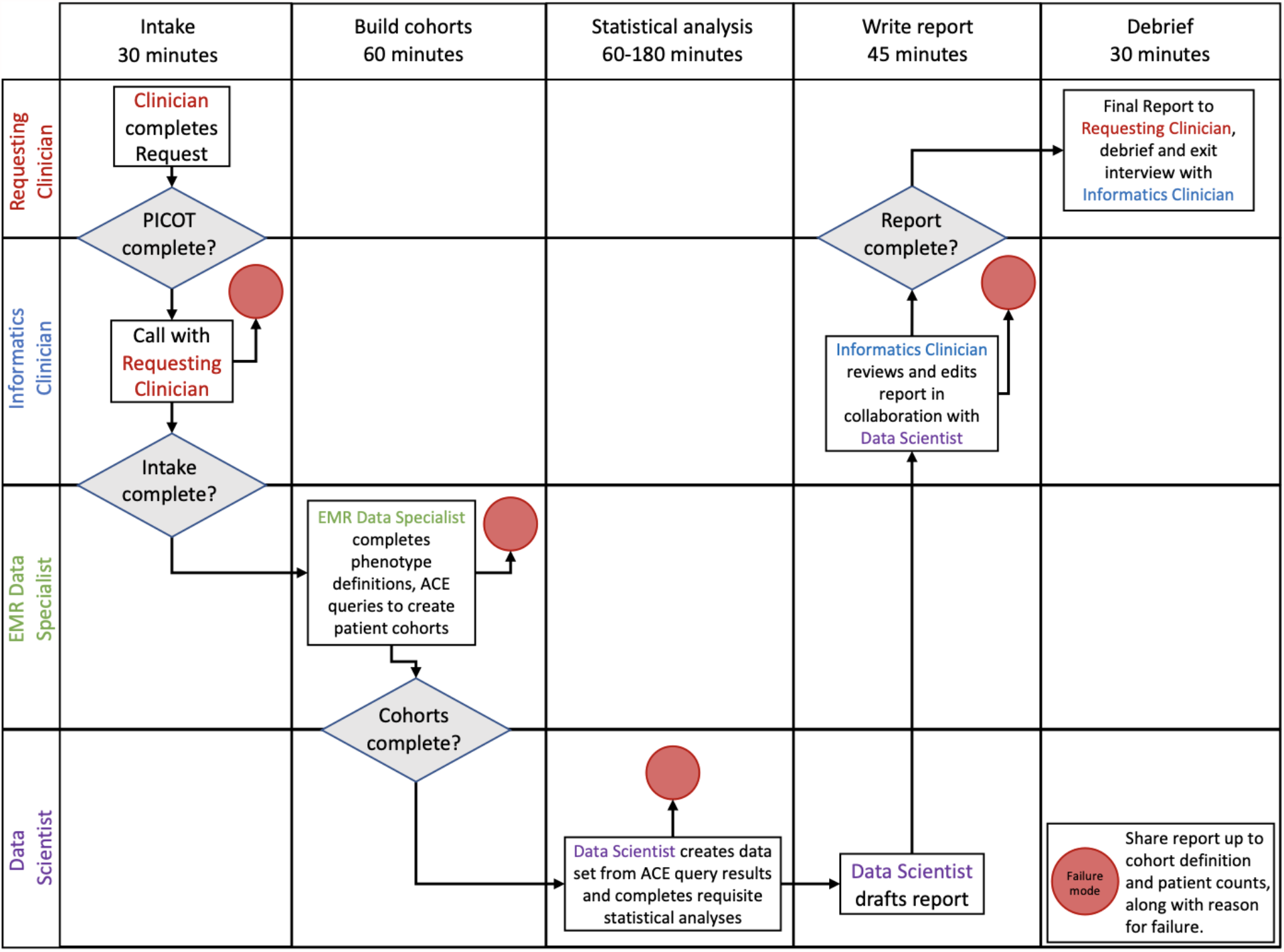
The workflow for fulfilling a consultation request, illustrating the order of each step, the time required, and the personnel responsible.

### Datasets and Cohort Building

The service used demographics, diagnoses, procedures, medications, laboratory values, clinical notes, length of stay, and mortality information for millions of patients from three data sources: EMRs from 3.1 million Stanford Medicine (*Stanford*) patients (54% female, spanning 1995-2019)^39^ consisting of diagnosis, procedure, medication, and laboratory test records, as well as clinical notes processed using a previously developed and evaluated text-processing pipeline^40,41^ ; IBM MarketScan® (*MarketScan*) which contains employer and Medicare insurance claims for 124 million lives (53% female, spanning 2007-2015); and Optum Clinformatics Data Mart® (*Optum*) which contains insurance claims for 53 million lives from employer sponsored health plans (53% female, spanning 2003-2016).

The choice of dataset for a given consultation was informed by the question and primarily based on meeting the criteria specified in the PICOT. For example, if a patient cohort definition relied on a specific range of laboratory test result values, then this necessitated using the Stanford EMR dataset, because claims data do not include laboratory test results. The EMR data specialist constructed patient cohorts using the Advanced Cohort Engine (ACE)^18^ to define necessary and sufficient conditions to determine if an exposure or outcome of interest occurred in a patient’s timeline. A patient timeline view of patient records provided by ACE enabled anonymized chart review for quality checks of the exposure and outcome definitions and resulting cohorts.

### Supported Analyses

The service supported treatment comparisons for discrete, continuous and time-to-event outcomes as well as custom descriptive analyses^16^. For discrete, continuous and time-to-event outcomes we used a standardized process which attempted to emulate a “target trial”^42^ based on the criteria specified in the PICOT. For consultations requesting treatment comparisons, we created cohorts of similar patients using two approaches: Mahalanobis distance with a fixed caliper based on age, gender, length of record, and year of entry into the cohort (we call this “simple matching”) or high dimensional propensity score matching (hd-PSM)^43,44^. Matching is a way to identify subsets of patients that are similar in most respects other than the treatment they received, in order to reduce the chance that observed differences in outcomes are due to variation in properties other than treatment but which also impact the outcome (commonly referred to as confounding)^43^. For propensity score estimation, we used an L2 regularized logistic regression model with a time-binned count based featurization of pre-treatment clinical data elements (diagnoses, procedures, medication records), fit using GLMnet^45^. Regularization strength was determined using 10-fold cross validation with a final refit on the entire data before estimating propensity scores for all patients^46^. Results from both matching strategies were included with each report.

The subsequent analysis performed on matched cohorts was selected based on the outcome specified in the PICOT formulation of the question. For treatment comparisons with binary outcomes we calculated odds ratios and associated confidence intervals. For treatment comparisons with continuous outcomes, we fit regression models and reported mean change in response estimates and associated confidence intervals. For treatment comparisons with time-to-event (survival) outcomes, we computed Kaplan-Meier plots and performed log rank tests for differences in survival curves between compared treatments, and reported hazard ratios (HRs) and associated confidence intervals.

Custom descriptive analyses required bespoke code for each request, primarily written in R, with data aggregation using Python as necessary. All analyses were conducted in R. Analysis code is publicly available^47^.

### Quality checks for supported treatment comparison analyses

Given that there is no known ground truth for the questions received by the consultation service, we established code correctness using synthetic datasets as well as derived an estimate of the false positive rate for treatment comparison analyses using publicly available datasets of drug-effect pairs as ground truth.

#### Establishing code correctness

We generated eight synthetic datasets, each with 10,000 patients, using all combinatorial variations of three properties: whether a binary treatment had an effect on a single survival outcome, whether treatment assignment had a dependence on a single binary covariate, and whether the covariate had an effect on the survival outcome. We confirmed the correctness of the analysis code by verifying that the analyses returned a treatment effect if and only if the underlying data were constructed with a treatment effect, and that the direction of the derived treatment effect was concordant with the treatment effect specified when creating the synthetic dataset. On performing treatment comparisons using cohorts matched with hd-PSM, the analysis code correctly identified protective effects for the four synthetic datasets constructed to have such intervention effects and no effects for the four synthetic datasets constructed to have no effect. For the two synthetic datasets where there was both a biased treatment and a covariate effect, resulting in confounding, propensity matching correctly recovered the true effect for the treatment.

#### Quantifying the expected false positive rate

We selected treatment-outcome pairs known to be either associated or non-associated as compiled by the OMOP community (Ryan et al^48^, 399 pairs) and the EU-ADR project (Coloma et al^20^, 93 pairs) because they are publicly available and have been used as ground truth sets in other studies^49–51^. While both reference sets contain known associations as well as asserted non-associations, only the asserted non-associations were informative in quantifying the false positive rate. Of the 399 drug-outcome pairs from the OMOP community reference set, 234 are non-associations. Of the 93 pairs from the EU-ADR project, 50 are non-associations.

We constructed cohorts corresponding to each of the asserted non-associated treatment-outcome pairs in the reference sets and estimated a treatment effect, using Stanford EMR data. Cohorts were constructed by transforming each treatment and outcome definition into corresponding ACE queries. Outcomes were defined using ICD9 codes and drug treatments were defined using RxNorm codes. We used a new patient cohort design where patients entered a cohort immediately after the first time they were prescribed a drug. Outcomes were measured as events after the first prescription, with patients being marked as censored when their medical records ended. A result was counted as false positive if our analysis found that a given treatment was associated with an increased or decreased hazard ratio relative to the comparator (with an effect estimate greater than or less than 1, and a p-value ≤ 0.05), and the reference set marked it as not associated.

Of the 234 non-associated pairs from the OMOP community, there were 137 drug-outcome pairs for which a minimum 100 patients exposed to the drug were present in Stanford data. Of these, 27 associations were false positives and the remaining 110 were correctly identified as non-associations, providing an estimated false positive rate of 20%. From the 50 non-associated treatment-outcome pairs from the EU-ADR project, there were 42 pairs for which there was enough data. Of these, 7 associations were false positives and the remaining 35 correctly identified as non-associations, providing an estimated false positive rate of 17%.

Because the OMOP and EU-ADR reference sets were constructed to evaluate methods for treatment comparisons, the 17-20% expected false positive rate is applicable to consultations requesting a comparison of the hazard ratio of an outcome between treatments.

## Summarizing the first 100 consultation requests

### Categorizing motivations for requests and subsequent actions

We categorized the scenarios motivating consultation requests, and subsequent actions by requestors, based on the intake and debrief meetings, respectively. Each consultation request was assigned a single motivation category, and one or more subsequent action categories. We categorized subsequent actions into one or more of three possible categories. If, during debrief, the requestor stated that they would use the knowledge gained from the consultation to change the treatment of a current or future patient with similar presentation, the consultation was categorized as having *changed patient care*. Debriefs where the requestor planned to obtain approval to further study their question or use the findings from the consultation to generate hypotheses for an ongoing research project were categorized as *guiding further research*. Debriefs where the requester used the results from the consultation report directly as the basis of a publication, poster, abstract, grant submission, or presentation were categorized as *follow-up analyses*. Because the motivating scenarios were not known in advance, the eight categories of motivation (Table 2) were developed after the 100 consultations were completed.

### Concordance of consultation results across data sources

We compared results obtained using different data sources for the same consultation request. To do so, we first identified consultations requesting treatment effect comparisons which could be re-executed using another dataset. For example, if a consultation was originally completed using data from Stanford, we re-executed it using MarketScan and Optum claims data. Some two-way comparisons across datasets failed due to few patients in a given dataset (our threshold was 100 patients), while for others the matching procedure resulted in groups with no overlap in their propensity score distributions and thus were unsuitable for comparison^52^.

Because a consultation to provide a treatment comparison could involve more than one outcome, we summarized concordance in terms of the number of outcomes, rather than the number of consultations. We evaluated the concordance of results for 59 outcomes from 33 consultations across Stanford and Optum; and 53 outcomes from 22 consultations across Stanford and MarketScan.

Using the notion of ‘regulatory agreement’^53^, a result was counted as concordant across two datasets only if both datasets provided an effect estimate in the same direction (e.g. both greater than 1 or both less than 1) with a p-value ≤ 0.05, or if the effect estimates derived from both datasets did not indicate a significant effect on the outcome(s) of interest, regardless of direction.

## Findings from the first 100 consultations

### Consultations requests came from multiple specialties

Of the first 100 requests by 53 users from multiple specialties, 83 consultations were completed. 17 consultations could not be completed due to missing data elements, available data sources having too few patients meeting the specified cohort criteria, inability to define a cohort, or requiring an unsupported study design.

Of the 83 completed consultations, 48 were descriptive analyses. 35 were treatment comparison analyses, of which 18 had discrete or continuous outcomes and 17 had time-to-event outcomes. 78 out of 83 (94%) consultations used Stanford EMR data, and 4 out of 83 (5%) used national claims data to obtain adequate sample size. One consultation used both EMR and claims data.

Internal medicine was the most common requesting specialty, in terms of both requests received and number of requestors, followed by dermatology, oncology and cardiology (Table 1). Among 53 users, 24 requested a consultation more than once, for a total of 76 consultations. Internal medicine also had the highest number of repeat users.

**Table 1.** Summary of completed consultations by specialty, showing the number of consultations of each analysis type.

| Specialty | Consultations | Analysis type |  |  |
| --- | --- | --- | --- | --- |
|  |  | Descriptive | Treatment comparison |  |
|  |  |  | Discrete or Continuous | Time-to-event |
| Internal Medicine | 21 | 10 | 11 | 0 |
| Dermatology | 9 | 8 | 1 | 0 |
| Oncology | 9 | 1 | 1 | 7 |
| Cardiology | 6 | 1 | 0 | 5 |
| General Pediatrics | 5 | 5 | 0 | 0 |
| Emergency Medicine | 4 | 2 | 2 | 0 |
| Endocrinology | 4 | 2 | 0 | 2 |
| Hematology | 4 | 1 | 1 | 2 |
| Allergy and Immunology | 3 | 3 | 0 | 0 |
| Infectious Disease | 3 | 3 | 0 | 0 |
| Pediatric Neurology | 3 | 3 | 0 | 0 |
| Vascular Surgery | 3 | 1 | 2 | 0 |
| Anesthesiology | 2 | 2 | 0 | 0 |
| Orthopaedic surgery | 2 | 2 | 0 | 0 |
| Pathology | 2 | 2 | 0 | 0 |
| Nephrology | 1 | 1 | 0 | 0 |
| Ophthalmology | 1 | 0 | 0 | 1 |
| Urology | 1 | 1 | 0 | 0 |
| **Total** | **83** | **48** | **18** | **17** |

Median consultation turnaround time was 5 days, with 71 (86%) of consultations completed in 10 days or less. Longer turnaround times occurred when additional data elements were needed, there were delays in scheduling conversations with the requestor, or when matching required substantial time for large cohorts. As the service workflow matured, by the end of the study, 19 consultation reports were returned in 48 hours or less by reusing cohort definitions, experience in PICOT formulation of the request, and analysis code optimization.

### Consultation requests had diverse motivations

Consultation requests were driven by a variety of motivations, including evaluating patient management strategies for a given disease or patient presentation, identifying comparatively effective treatments for patients with typically understudied characteristics, and quantifying associations between diseases. The categorization of consultation motivations is summarized in Table 2, and cross-tabulated with the subsequent actions taken by requestors. 10 consultations led to changes to patient care, 52 guided further research, and 17 led to follow up analyses, including four that were presented at medical conferences or published in peer-reviewed journals^54–57^. Not all subsequent actions could be categorized into the three groups: 27 consultations lacked clear subsequent actions, suggesting that the consultation may have been sought primarily to contribute to the personal knowledge of the requestor, or that the findings were not sufficiently compelling to warrant action on their basis.

**Table 2.**
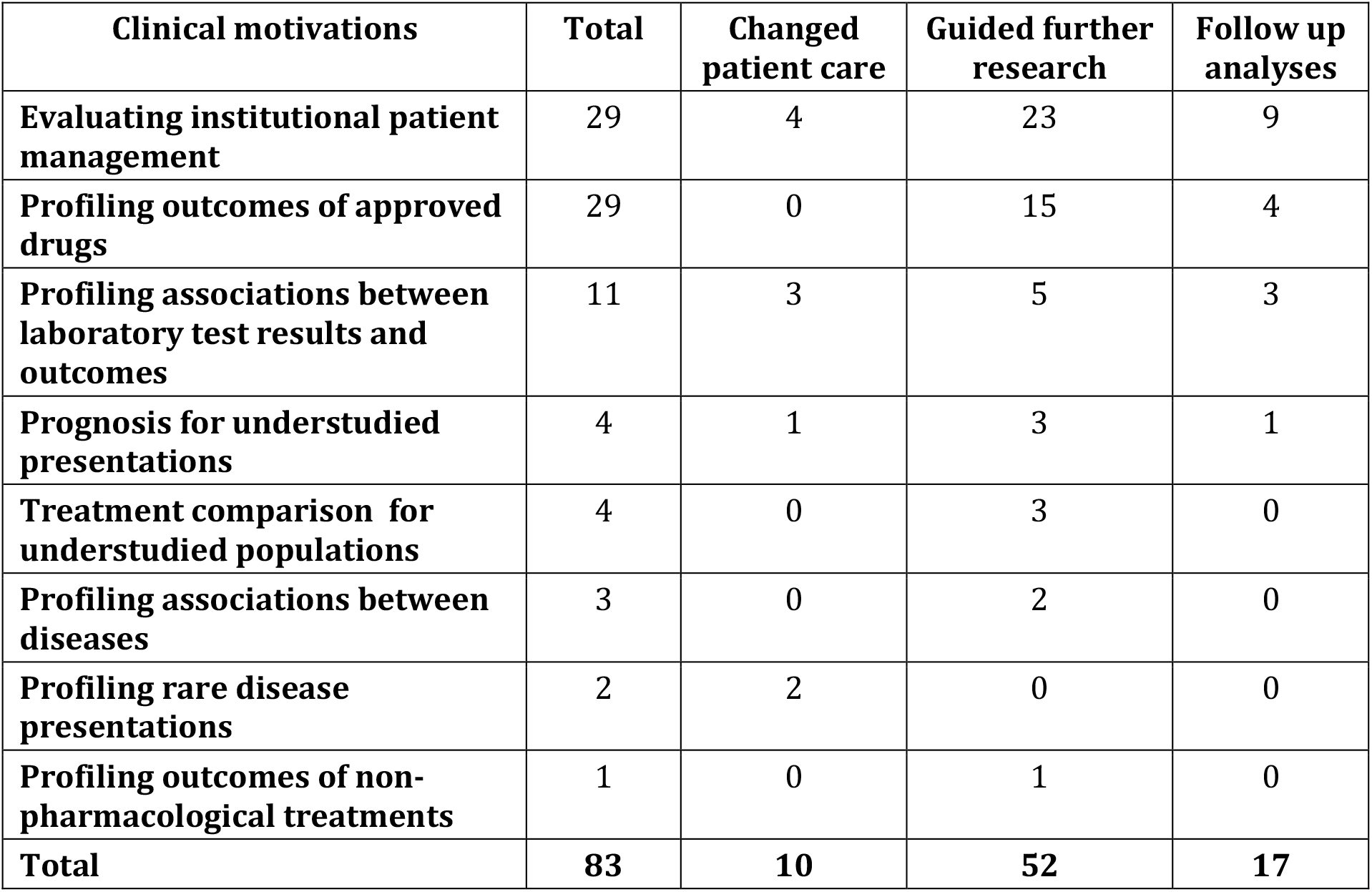
Clinical motivations and subsequent actions taken by requestors for the 83 completed consultations. Each request was assigned a single motivation category and one or more follow-up action categories.

We highlight three consultations that demonstrate the diversity of situations motivating a consultation: a request to characterize a rare disease presentation (a pediatric patient with mononeuritis multiplex); a request to compare treatment outcomes (for a recently approved class of melanoma drugs, PD-1 inhibitors); and a request to summarize the institutional use of procalcitonin tests (to inform antibiotic discontinuation). The reports for these three consultations are provided in the Supplement. In each of these consultations, the service addressed a different need.

In the case of the pediatric mononeuritis multiplex patient the consultation required a custom descriptive analysis of a rare disease presentation that resulted in changes to patient care. We provided the requestor with summaries of the most frequent diagnoses preceding and following mononeuritis multiplex diagnosis in 118 similarly aged patients, which included bacterial and viral infections as well as psychosomatic disorders. A variety of treatments were prescribed for those patients, including steroids, antibiotics, anti-inflammatory medications, painkillers, and hormone supplements. These findings, alongside further clinical workup suggested managing the patient’s symptoms as a post-viral syndrome. The patient improved with a trial of steroids and was discharged.

In the case of the use of PD-1 inhibitors the consultation required a treatment comparison analysis for an understudied population that guided further research. The consultation was motivated by a melanoma patient who had a herpes simplex reactivation following treatment with nivolumab. We found 587 similar patients and found no difference in viral reactivation rates in patients treated with PD-1 inhibitors compared to those treated with other antineoplastic agents. Published evidence on the relationship between PD-1 therapies and herpetic reactivations was not available, perhaps because nivolumab was only recently approved (in 2014). Here, the consultation findings filled an important clinical evidence gap.

In the case of procalcitonin testing, the request entailed a custom descriptive analysis to evaluate institutional patient management that changed patient care. The consultation was requested because while procalcitonin is a serum biomarker capable of discriminating between bacterial and non-bacterial causes of infection^58–61^ the exact cut-off value at which to discontinue antibiotics is not universally agreed upon. Procalcitonin’s utility for deciding whether to order a blood culture remains unclear^62,63^. By analyzing approximately 16,000 procalcitonin test results and 29,000 blood culture results, we calculated how often a positive blood culture was obtained within 48 hours of one cut-off value for a procalcitonin result, how frequently antibiotic therapy was discontinued at different cut-offs of procalcitonin values, and how often antibiotics were restarted within 72 hours of discontinuation. The analysis found that at the cut-offs in use (procalcitonin > 0.5) a positive test was not associated with a positive blood culture. This finding, combined with further analyses, informed an institutional protocol change: procalcitonin values are no longer used to inform ordering a blood culture when deciding whether to discontinue antibiotic therapy.

### Consultation results were concordant across datasets

When comparing results obtained using different data sources for the same consultation request, 68% to 74% of results were concordant across datasets. In the Stanford and Optum comparison, results for 68% (40 out of 59) of the evaluated outcomes were concordant. For 28 outcomes, both datasets reported a significant treatment effect with the same direction of the effect. For 12 outcomes, results from both datasets did not have a significant effect. In the Stanford and MarketScan comparison, 74% (39 out of 53) of the evaluated outcomes were concordant. For 30 outcomes, results from both datasets had a significant treatment effect with the same direction of the effect. For 9 outcomes, results from both datasets did not have a significant effect.

## A vision realized: strengths, caveats and next steps

Using data generated during routine care to guide the care of future patients is a core tenet of a learning health system^64–67^ and, as a distillation of clinical expertise, of evidence-based medicine^1,68^. Our work is a first-of-its-kind implementation of this vision, demonstrating that an on-demand consultation service to summarize the experiences of previously seen patients is feasible from both an engineering and operational standpoint. The variety of consultation requests we received (in terms of clinical motivations, analyses needed and subsequent actions) also empirically illustrates the potential to inform a broad range of clinical decisions (Figure 2). As large patient data repositories become commonplace ^75,76^, the ability to learn from the experience of similar patients is one of the nobler opportunities such repositories enable^77^.

**Figure 2.**
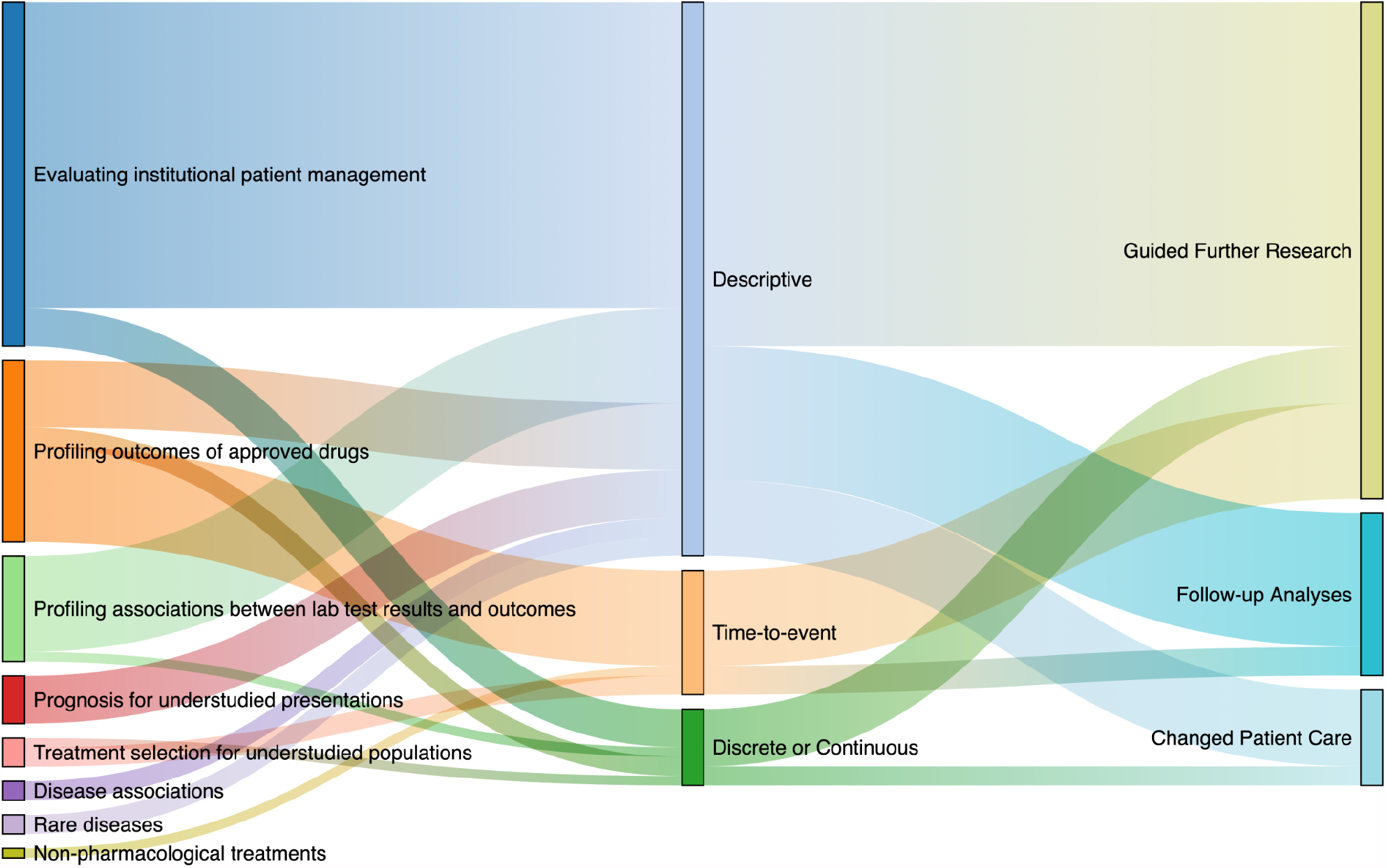
Sankey plot illustrates the flow (horizontal colored lines) of completed consultations in terms of the clinical motivation (left) to the analysis type (center) to the subsequent action (right). The thickness of each flow is proportional to the number of consultations. For example, consultations motivated by evaluating institutional patient management required mostly descriptive analyses, and resulted in all three categories of subsequent action.

Our work has several unique strengths. First, the service’s underlying search engine, ACE—which is essential for rapidly constructing cohorts and defining electronic phenotypes corresponding to exposures and outcomes of interest—is freely available for non-commercial use, allowing implementation of such a service at other sites without extensive monetary or technical resources^18^. Second, we found 68-74% concordance of consultation results across multiple datasets, a rate of agreement comparable both to the rate at which results from randomized controlled trials (RCTs) agree with each other (67-87%)^69^ and to the rate at which results derived from observational claims data agree with RCTs (60-80%)^70^. Finally, the number of repeat requests demonstrates the need for, and viability of, such a consultation service.

Our study has several limitations. First, users of the service were self-selecting; consultation requestors may thus have been predisposed to finding value in the service, and self-reported utility for advancing research or patient care may have been affected by subjective expectations of the service. Second, the cost to deploy such a service will vary at institutions where the necessary data access and analysis infrastructure does not yet exist. Our implementation was at an academic medical center with ready access to EMR and claims data resulting in an estimated cost per consult at $550 USD^14,16^ ; costs may be higher elsewhere. Moreover, while the current turnaround time is analogous to a send-out laboratory test, providing an ongoing service would require additional engineering effort incurring additional costs^71^. Third, the choice and evaluation of patient matching and causal inference methods remains an active area of research^72–74^. Future work may find methods beyond hd-PSM that offer improved concordance across data sources. Lastly, the net-benefit of providing on-demand evidence needs to be studied prospectively at multiple sites by measuring impact on patient outcomes, cost of care, and health system operations. We hope that our experience and the tooling we share will enable such studies.

## Takeaways

On-demand evidence generation to inform clinical decision making is an achievable goal, given the confluence of scalable technology for data analysis, a growing data science workforce, the training of increasingly data savvy clinicians, and the availability of large amounts of patient data from EMRs and claims ^8,14^. The consultation service we created provides proof-of-feasibility for realizing this goal. Such a service is capable of informing patient care at the bedside for specific patients, informing the medical literature and supporting institutional guideline creation. As large patient data repositories are created^75,76^, the potential to benefit from such a service is immense^77^. Given the feasibility, and the documented need, studies establishing the utility of having such a consultation service are logical next steps.

## Data Availability

Data are available upon reasonable request.

## References

1. Sackett DL, Rosenberg WM, Gray JA, Haynes RB, Richardson WS. Evidence based medicine: what it is and what it isn’t. BMJ 1996;312(7023):71–2.

2. Institute of Medicine (US) Committee on Quality of Health Care in America. To Err is Human: Building a Safer Health System. Washington (DC): National Academies Press (US); 2014.

3. Makary MA, Daniel M. Medical error-the third leading cause of death in the US. BMJ. 2016;353:i2139.

4. Rogers JR, Liu C, Hripcsak G, Cheung YK, Weng C. Comparison of Clinical Characteristics Between Clinical Trial Participants and Nonparticipants Using Electronic Health Record Data. JAMA Netw Open 2021;4(4):e214732.

5. Black N. Why we need observational studies to evaluate the effectiveness of health care. BMJ 1996;312(7040):1215–8.

6. Stewart WF, Shah NR, Selna MJ, Paulus RA, Walker JM. Bridging the inferential gap: the electronic health record and clinical evidence. Health Aff 2007;26(2):w181–91.

7. Califf RM, Robb MA, Bindman AB, et al. Transforming Evidence Generation to Support Health and Health Care Decisions. N Engl J Med 2016;375(24):2395–400.

8. Longhurst CA, Harrington RA, Shah NH. A “green button” for using aggregate patient data at the point of care. Health Aff 2014;33(7):1229–35.

9. Institute of Medicine, Roundtable on Evidence-Based Medicine. The Learning Healthcare System: Workshop Summary. National Academies Press; 2007.

10. Feinstein AR, Rubinstein JF, Ramshaw WA. Estimating prognosis with the aid of a conversational-mode computer program. Ann Intern Med 1972;76(6):911–21.

11. Rosati RA, McNeer JF, Starmer CF, Mittler BS, Morris JJ Jr, Wallace AG. A new information system for medical practice. Arch. Intern. Med. 1975;135(8):1017–24.

12. Frankovich J, Longhurst CA, Sutherland SM. Evidence-based medicine in the EMR era. N Engl J Med 2011;365(19):1758–9.

13. Institute of Medicine, Roundtable on Value and Science-Driven Health Care. Digital Infrastructure for the Learning Health System: The Foundation for Continuous Improvement in Health and Health Care: Workshop Series Summary. National Academies Press; 2011.

14. Gombar S, Callahan A, Califf R, Harrington R, Shah NH. It is time to learn from patients like mine. npj Digital Medicine. 2019;2:16.

15. Callahan A, Shah NH, Chen JH. Research and Reporting Considerations for Observational Studies Using Electronic Health Record Data. Ann Intern Med 2020;172(11 Suppl):S79–84.

16. Schuler A, Callahan A, Jung K, Shah NH. Performing an Informatics Consult: Methods and Challenges. J Am Coll Radiol 2018;15(3 Pt B):563–8.

17. Datta S, Posada J, Olson G, et al. A new paradigm for accelerating clinical data science at Stanford Medicine. arXiv [cs.CY]. 2020; http://arxiv.org/abs/2003.10534.

18. Callahan A, Polony V, Posada JD, Banda JM, Gombar S, Shah NH. ACE: the Advanced Cohort Engine for searching longitudinal patient records. J Am Med Inform Assoc 2021; http://dx.doi.org/10.1093/jamia/ocab027.10.1093/jamia/ocab027.

19. Wang G, Jung K, Winnenburg R, Shah NH. A method for systematic discovery of adverse drug events from clinical notes. J Am Med Inform Assoc 2015;22(6):1196–204.

20. LePendu P, Iyer SV, Bauer-Mehren A, et al. Pharmacovigilance using clinical notes. Clin Pharmacol Ther 2013;93(6):547–55.

21. Callahan A, Fries JA, Ré C, et al. Medical device surveillance with electronic health records. NPJ Digit Med 2019;2:94.

22. Nead KT, Gaskin G, Chester C, et al. Androgen Deprivation Therapy and Future Alzheimer’s Disease Risk. J Clin Oncol 2016;34(6):566–71.

23. Nead KT, Gaskin G, Chester C, Swisher-McClure S, Leeper NJ, Shah NH. Association Between Androgen Deprivation Therapy and Risk of Dementia. JAMA Oncol 2017;3(1):49–55.

24. Tamang S, Patel MI, Blayney DW, et al. Detecting unplanned care from clinician notes in electronic health records. J Oncol Pract 2015;11(3):e313–9.

25. Hernandez-Boussard T, Tamang S, Blayney D, Brooks J, Shah N. New Paradigms for Patient-Centered Outcomes Research in Electronic Medical Records: An Example of Detecting Urinary Incontinence Following Prostatectomy. EGEMS (Wash DC) 2016;4(3):1231.

26. Caswell-Jin JL, Callahan A, Purington N, et al. Treatment and Monitoring Variability in US Metastatic Breast Cancer Care. JCO Clinical Cancer Informatics (2021);5:600–614.

27. Vashisht R, Jung K, Schuler A, et al. Association of Hemoglobin A1c Levels With Use of Sulfonylureas, Dipeptidyl Peptidase 4 Inhibitors, and Thiazolidinediones in Patients With Type 2 Diabetes Treated With Metformin: Analysis From the Observational Health Data Sciences and Informatics Initiative. JAMA Netw Open 2018;1(4):e181755.

28. Banda JM, Seneviratne M, Hernandez-Boussard T, Shah NH. Advances in Electronic Phenotyping: From Rule-Based Definitions to Machine Learning Models. Annu Rev Biomed Data Sci 2018;1:53–68.

29. Seneviratne MG, Banda JM, Brooks JD, Shah NH, Hernandez-Boussard TM. Identifying Cases of Metastatic Prostate Cancer Using Machine Learning on Electronic Health Records. AMIA Annu Symp Proc 2018;2018:1498–504.

30. Agarwal V, Podchiyska T, Banda JM, et al. Learning statistical models of phenotypes using noisy labeled training data. J Am Med Inform Assoc 2016;23(6):1166–73.

31. Banda JM, Halpern Y, Sontag D, Shah NH. Electronic phenotyping with APHRODITE and the Observational Health Sciences and Informatics (OHDSI) data network. AMIA Jt Summits Transl Sci Proc 2017;2017:48–57.

32. Kim Y, Tian Y, Yang J, et al. Comparative safety and effectiveness of alendronate versus raloxifene in women with osteoporosis. Sci Rep 2020;10(1):11115.

33. Chen R, Ryan P, Natarajan K, et al. Treatment Patterns for Chronic Comorbid Conditions in Patients With Cancer Using a Large-Scale Observational Data Network. JCO Clin Cancer Inform 2020;4:171–83.

34. Prats-Uribe A, Sena AG, Lai LYH, et al. Use of repurposed and adjuvant drugs in hospital patients with covid-19: multinational network cohort study. BMJ 2021;373:1038.

35. Tan EH, Sena AG, Prats-Uribe A, et al. COVID-19 in patients with autoimmune diseases: characteristics and outcomes in a multinational network of cohorts across three countries. Rheumatology 2021; http://dx.doi.org/10.1093/rheumatology/keab250.

36. Burn E, You SC, Sena AG, et al. Deep phenotyping of 34,128 adult patients hospitalised with COVID-19 in an international network study. Nat Commun 2020;11(1):5009.

37. Duke JD, Ryan PB, Suchard MA, et al. Risk of angioedema associated with levetiracetam compared with phenytoin: Findings of the observational health data sciences and informatics research network. Epilepsia 2017;58(8):e101–6.

38. Hripcsak G, Ryan PB, Duke JD, et al. Characterizing treatment pathways at scale using the OHDSI network. Proc Natl Acad Sci U S A 2016;113(27):7329–36.

39. Lowe HJ, Ferris TA, Hernandez PM, Weber SC. STRIDE--An integrated standards-based translational research informatics platform. AMIA Annu Symp Proc 2009;2009:391–5.

40. Lependu P, Iyer SV, Fairon C, Shah NH. Annotation Analysis for Testing Drug Safety Signals using Unstructured Clinical Notes. J Biomed Semantics 2012;3 Suppl 1:S5.

41. Lependu P, Liu Y, Iyer S, Udell MR, Shah NH. Analyzing patterns of drug use in clinical notes for patient safety. AMIA Jt Summits Transl Sci Proc 2012;2012:63–70.

42. Hernán MA, Robins JM. Using Big Data to Emulate a Target Trial When a Randomized Trial Is Not Available. Am J Epidemiol 2016;183(8):758–64.

43. Austin PC. An Introduction to Propensity Score Methods for Reducing the Effects of Confounding in Observational Studies. Multivariate Behav Res 2011;46(3):399–424.

44. Guertin JR, Rahme E, Dormuth CR, LeLorier J. Head to head comparison of the propensity score and the high-dimensional propensity score matching methods. BMC Med Res Methodol 2016;16:22.

45. Friedman J, Hastie T, Tibshirani R. Regularization Paths for Generalized Linear Models via Coordinate Descent. J Stat Softw 2010;33(1):1–22.

46. Stuart EA. Matching methods for causal inference: A review and a look forward. Stat Sci 2010;25(1):1–21.

47. Steinberg E, Callahan A, Shah NH. Green Button analysis code. 2020; Accessed May 28, 2021. https://github.com/som-shahlab/green_button.

48. Coloma PM, Avillach P, Salvo F, et al. A reference standard for evaluation of methods for drug safety signal detection using electronic healthcare record databases. Drug Saf 2013;36(1):13–23.

49. Eriksson R, Werge T, Jensen LJ, Brunak S. Dose-specific adverse drug reaction identification in electronic patient records: temporal data mining in an inpatient psychiatric population. Drug Saf 2014;37(4):237–47.

50. Xiao C, Li Y, Baytas IM, Zhou J, Wang F. An MCEM Framework for Drug Safety Signal Detection and Combination from Heterogeneous Real World Evidence. Sci Rep 2018;8(1):1806.

51. Ryan PB, Buse JB, Schuemie MJ, et al. Comparative effectiveness of canagliflozin, SGLT2 inhibitors and non-SGLT2 inhibitors on the risk of hospitalization for heart failure and amputation in patients with type 2 diabetes mellitus: A real-world meta-analysis of 4 observational databases (OBSERVE-4D). Diabetes Obes Metab 2018;20(11):2585–97.

52. Oberst M, Johansson FD, Wei D, et al. Characterization of Overlap in Observational Studies. arXiv [cs.LG]. 2019; http://arxiv.org/abs/1907.04138.

53. Franklin JM, Pawar A, Martin D, et al. Nonrandomized Real-World Evidence to Support Regulatory Decision Making: Process for a Randomized Trial Replication Project. Clin Pharmacol Ther 2019; http://dx.doi.org/10.1002/cpt.1633.10.1002/cpt.1633.

54. Rhee J, Gombar S, Beutner K, Callahan A, Jung K, Wheeler M. Cardiovascular safety of mexiletine as a therapy for myotonia in patients with myotonic dystrophy. J Am Coll Cardiol 2019;73(9):474.

55. Karimi Y, Gombar S, Dean L, et al. Real-world efficacy of bone modifying agents (BMAs) in patients with breast cancer (BC) treated in an academic health system: Use of the “green button. J Clin Orthod 2019;37(15_suppl):e18054–e18054.

56. Meng L, Gombar S, Callahan A, et al. 210. Step-down from IV to oral therapy in patients with bacteremia due to Enterobacteriaceae: fluoroquinolones (FQ) vs. ß-lactams (BL) or trimethoprim-sulfamethoxazole (TMP-SMX). Open Forum Infectious Diseases. 2019;6(Supplement_2):S124–S124. http://dx.doi.org/10.1093/ofid/ofz360.285.

57. Ibrahim B, de Freitas Mendonca MI, Gombar S, Callahan A, Jung K, Capasso R. Association of Systemic Diseases With Surgical Treatment for Obstructive Sleep Apnea Compared With Continuous Positive Airway Pressure. JAMA Otolaryngol Head Neck Surg 2021; http://dx.doi.org/10.1001/jamaoto.2020.5179.10.1001/jamaoto.2020.5179.

58. Christ-Crain M, Jaccard-Stolz D, Bingisser R, et al. Effect of procalcitonin-guided treatment on antibiotic use and outcome in lower respiratory tract infections: cluster-randomised, single-blinded intervention trial. Lancet 2004;363(9409):600–7.

59. Schuetz P, Christ-Crain M, Thomann R, et al. Effect of procalcitonin-based guidelines vs standard guidelines on antibiotic use in lower respiratory tract infections: the ProHOSP randomized controlled trial. JAMA 2009;302(10):1059–66.

60. Tonkin-Crine SK, Tan PS, van Hecke O, et al. Clinician-targeted interventions to influence antibiotic prescribing behaviour for acute respiratory infections in primary care: an overview of systematic reviews. Cochrane Database Syst Rev 2017;9:CD012252.

61. Schuetz P, Wirz Y, Sager R, et al. Procalcitonin to initiate or discontinue antibiotics in acute respiratory tract infections. Cochrane Database Syst Rev 2017;10:CD007498.

62. Wu Q, Yang H, Kang Y. Comparison of diagnostic accuracy among procalcitonin, C-reactive protein, and interleukin 6 for blood culture positivity in general ICU patients. Crit Care 2018;22(1):339.

63. Bassetti M, Russo A, Righi E, et al. Comparison between procalcitonin and C-reactive protein to predict blood culture results in ICU patients. Crit Care 2018;22(1):252.

64. Grumbach K, Lucey CR, Johnston SC. Transforming from centers of learning to learning health systems: the challenge for academic health centers. JAMA 2014;311(11):1109–10.

65. Krumholz HM, Terry SF, Waldstreicher J. Data Acquisition, Curation, and Use for a Continuously Learning Health System. JAMA 2016;316(16):1669–70.

66. Smoyer WE, Embi PJ, Moffatt-Bruce S. Creating Local Learning Health Systems: Think Globally, Act Locally. JAMA 2016;316(23):2481–2.

67. Bindman AB, Pronovost PJ, Asch DA. Funding Innovation in a Learning Health Care System. JAMA 2018;319(2):119–20.

68. Evidence-Based Medicine Working Group. Evidence-based medicine. A new approach to teaching the practice of medicine. JAMA 1992;268(17):2420–5.

69. Ryan PB, Hripcsak G. A journey toward real-world evidence for regulatory decision-making: Proving reliable real-world evidence - Replicating RCTs using LEGEND. 2019; https://www.ohdsi.org/wp-content/uploads/2019/09/4-Plenary-3-Replicating-LEGEND.pdf.

70. Franklin JM, Patorno E, Desai RJ, et al. Emulating Randomized Clinical Trials With Nonrandomized Real-World Evidence Studies: First Results From the RCT DUPLICATE Initiative. Circulation 2021;143(10):1002–13.

71. Norgeot B, Glicksberg BS, Butte AJ. A call for deep-learning healthcare. Nat Med 2019;25(1):14–5.

72. King G, Lucas C, Nielsen RA. The balance-sample size frontier in matching methods for causal inference. Am J Pol Sci 2017;61(2):473–89.

73. Powers S, Qian J, Jung K, et al. Some methods for heterogeneous treatment effect estimation in high dimensions. Stat Med 2018;37(11):1767–87.

74. Liu L, Mukherjee R, Robins JM. On Nearly Assumption-Free Tests of Nominal Confidence Interval Coverage for Causal Parameters Estimated by Machine Learning. SSO Schweiz Monatsschr Zahnheilkd 2020;35(3):518–39.

75. Mandl KD, Kohane IS. Tectonic shifts in the health information economy. N Engl J Med 2008;358(16):1732–7.

76. Mandl KD, Perakslis ED. HIPAA and the Leak of “Deidentified” EHR Data. N Engl J Med 2021;384(23):2171–3.

77. Palmer K, Brodwin E, Sheridan K. The growing health data marketplace sparks concern over patient privacy. 2021; Accessed Jun 15, 2021. https://www.statnews.com/2021/06/09/datavant-ciox-health-data-hipaa/.

