## Supplement for "Using aggregate patient data at the bedside via an on-demand consultation service"

#### Example consultation reports

##### Example consultation report #1

**CLINICAL QUESTION**
In a young patient presenting with mononeuritis multiplex what is their ultimate diagnosis?

**HOW WE ASKED THIS QUESTION**
In patients aged ≤ 21 years and diagnosed with mononeuritis multiplex, with at least one year of data prior to and after initial diagnosis, what diagnoses were recorded in the 5 years prior to mononeuritis multiplex diagnosis? What diagnoses were recorded in the 5 years after mononeuritis multiplex diagnosis? What drugs were prescribed in the 5 years after mononeuritis multiplex diagnosis?

Data source: IBM Marketscan Database®

###### Researcher Interpretation

The 5 most observed diagnoses in the 5 years prior to mononeuritis multiplex diagnosis were acute pharyngitis (ICD9 462), acute upper respiratory infection (ICD9 465.9), pain in limb (ICD9 729.5), headache (ICD9 784) and acne (ICD9 706.1). The 5 most observed diagnoses in the 5 years following mononeuritis multiplex diagnosis were acute pharyngitis (ICD9 462), malaise and fatigue (ICD9 780.79), pain in limb (ICD9 729.5), acne (ICD9 706.1), and acute upper respiratory infection (ICD9 465.9). The 5 most prescribed medications in the 5 years following mononeuritis multiplex were azithromycin, amoxicillin, acetaminophen, hydrocodone and ethinyl estradiol.

###### Summary of Cohort

DEFINITIONS

1) **Mononeuritis multiplex:** ICD9 354.5

SUMMARY

118 patients had a diagnosis of mononeuritis multiplex when ≤ 21 years old, and had at least one year of data prior to and following diagnosis. Their demographics are summarized in Figure 1.

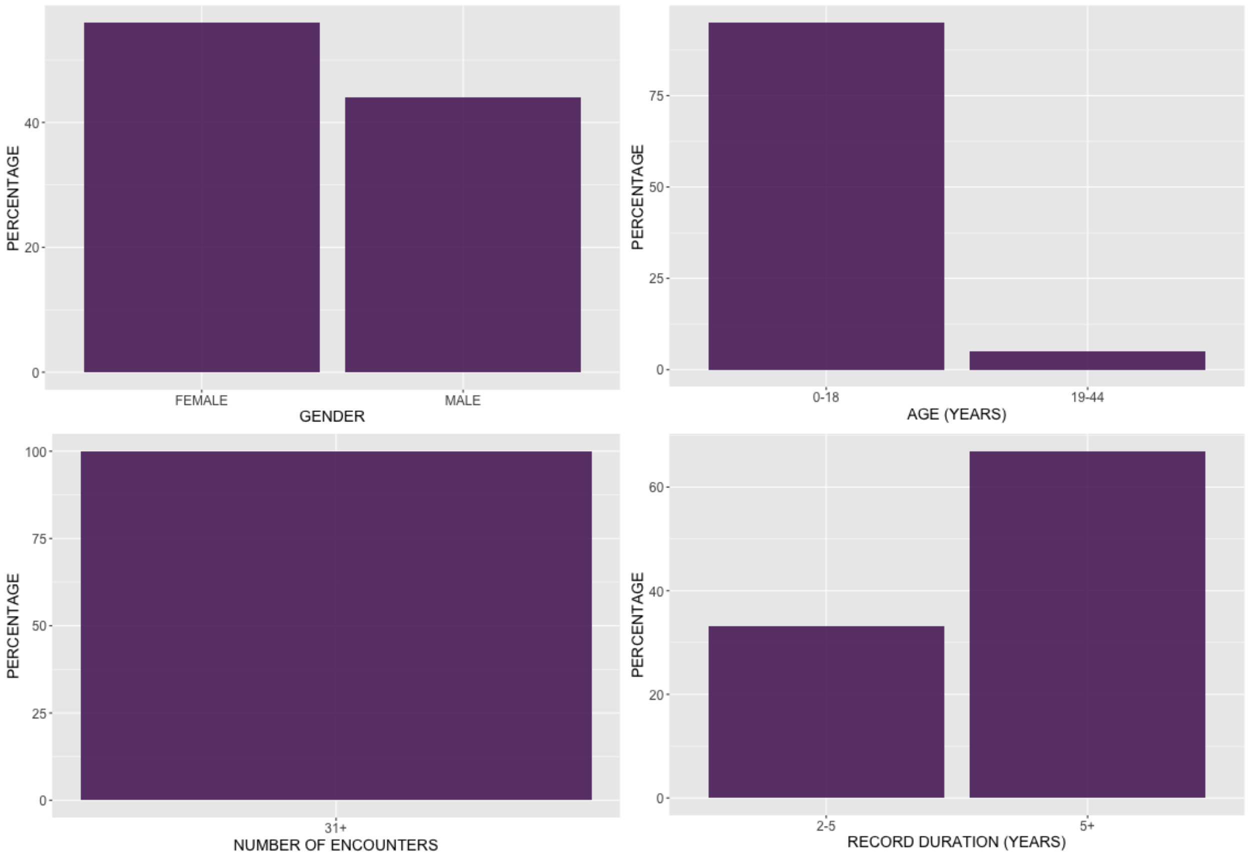

**Figure 1.** Demographics summary of patients ≤ 21 years old with a diagnosis of mononeuritis multiplex, and with at least one year of data prior and following diagnosis.

#### Research walkthrough

118 patients had a diagnosis of mononeuritis multiplex when ≤ 21 years old, and had at least one year of data prior to and following diagnosis. 61 unique diagnoses were observed in at least 10 patients in the 5 years prior to mononeuritis multiplex diagnosis (Table 1). 59 unique diagnoses were observed in at least 10 patients in the 5 years following mononeuritis multiplex diagnosis (Table 2). 32 unique drugs were prescribed to at least 10 patients in the 5 years following mononeuritis multiplex diagnosis (Table 3).

**Table 1.** Diagnoses in 5 years preceding mononeuritis multiplex diagnosis, observed in at least 10 patients.

| ICD9 | Name | # Patients | % Patients |
|---|---|---|---|
| 462 | acute pharyngitis | 53 | 44.92 |
| 465.9 | acute uri nos | 40 | 33.90 |
| 729.5 | pain in limb | 37 | 31.36 |
| 784 | headache | 32 | 27.12 |
| 706.1 | acne nec | 30 | 25.42 |
| 461.9 | acute sinusitis nos | 28 | 23.73 |
| 723.1 | cervicalgia | 28 | 23.73 |
| 780.79 | malaise and fatigue nec | 28 | 23.73 |
| 789 | abdmnal pain unspcf site | 28 | 23.73 |
| 786.2 | cough | 27 | 22.88 |
| 493.9 | asthma, unspecified type, unspecified | 25 | 21.19 |

| | | | |
|---|---|---|---|
| 477.9 | allergic rhinitis, cause unspecified | 24 | 20.34 |
| 782 | skin sensation disturb | 24 | 20.34 |
| 719.41 | joint pain-shlder | 23 | 19.49 |
| 719.46 | joint pain-l/leg | 23 | 19.49 |
| 79.99 | unspecified viral infection | 22 | 18.64 |
| 466 | acute bronchitis | 22 | 18.64 |
| 719.43 | joint pain-forearm | 19 | 16.10 |
| 719.47 | pain in joint, ankle and foot | 19 | 16.10 |
| 724.2 | lumbago | 18 | 15.25 |
| 355.9 | mononeuritis of unspecified site | 17 | 14.41 |
| 786.5 | chest pain nos | 17 | 14.41 |
| 723.4 | brachial neuritis nos | 16 | 13.56 |
| 780.4 | dizziness and giddiness | 16 | 13.56 |
| 787.02 | nausea alone | 16 | 13.56 |
| 382.9 | otitis media nos | 15 | 12.71 |
| 285.9 | anemia nos | 14 | 11.86 |
| 599 | urin tract infection nos | 14 | 11.86 |
| 719.42 | joint pain-up/arm | 14 | 11.86 |
| 724.5 | backache nos | 14 | 11.86 |
| 729.1 | myalgia and myositis nos | 14 | 11.86 |
| 782.1 | nonspecif skin erupt nec | 14 | 11.86 |
| 789.07 | abdmnal pain generalized | 14 | 11.86 |
| 34 | strep sore throat | 13 | 11.02 |
| 354.2 | ulnar nerve lesion | 13 | 11.02 |
| 780.2 | syncope and collapse | 13 | 11.02 |
| 787.91 | diarrhea | 13 | 11.02 |
| 959.3 | elb/forearm/wrst inj nos | 13 | 11.02 |
| 530.81 | esophageal reflux | 12 | 10.17 |
| 354 | carpal tunnel syndrome | 11 | 9.32 |
| 463 | acute tonsillitis | 11 | 9.32 |
| 473.9 | chronic sinusitis nos | 11 | 9.32 |
| 477 | rhinitis due to pollen | 11 | 9.32 |
| 692.9 | dermatitis nos | 11 | 9.32 |
| 719.44 | pain in joint, hand | 11 | 9.32 |
| 724.4 | lumbosacral neuritis nos | 11 | 9.32 |
| 729.2 | neuralgia/neuritis nos | 11 | 9.32 |
| 780.6 | fever nos | 11 | 9.32 |
| 786.09 | respiratory abnorm nec | 11 | 9.32 |
| 844.9 | sprain of knee & leg nos | 11 | 9.32 |
| 959.7 | lower leg injury nos | 11 | 9.32 |
| 88.81 | lyme disease | 10 | 8.47 |

| | | | |
|---|---|---|---|
| 311 | depressive disorder, not elsewhere classified | 10 | 8.47 |
| 353 | brachial plexus lesions | 10 | 8.47 |
| 558.9 | noninf gastroenterit nec | 10 | 8.47 |
| 564 | constipation nos | 10 | 8.47 |
| 728.87 | muscle weakness-general | 10 | 8.47 |
| 786.05 | shortness of breath | 10 | 8.47 |
| 842 | sprain of wrist, unspecified site | 10 | 8.47 |
| 845 | sprain of ankle, unspecified site | 10 | 8.47 |
| 959.01 | head injury, unspecified | 10 | 8.47 |

**Table 2.** Diagnoses in 5 years following mononeuritis multiplex diagnosis, observed in at least 10 patients.

| ICD9 | Name | # Patients | % Patients |
|---|---|---|---|
| 462 | acute pharyngitis | 52 | 44.07 |
| 780.79 | malaise and fatigue nec | 40 | 33.90 |
| 729.5 | pain in limb | 39 | 33.05 |
| 706.1 | acne nec | 37 | 31.36 |
| 465.9 | acute uri nos | 35 | 29.66 |
| 784 | headache | 31 | 26.27 |
| 461.9 | acute sinusitis nos | 27 | 22.88 |
| 789 | abdmnal pain unspcf site | 27 | 22.88 |
| 300 | anxiety state nos | 24 | 20.34 |
| 493.9 | asthma, unspecified type, unspecified | 24 | 20.34 |
| 782 | skin sensation disturb | 24 | 20.34 |
| 786.2 | cough | 23 | 19.49 |
| 466 | acute bronchitis | 22 | 18.64 |
| 723.1 | cervicalgia | 22 | 18.64 |
| 692.9 | dermatitis nos | 21 | 17.80 |
| 729.1 | myalgia and myositis nos | 21 | 17.80 |
| 786.5 | chest pain nos | 20 | 16.95 |
| 599 | urin tract infection nos | 18 | 15.25 |
| 728.87 | muscle weakness-general | 18 | 15.25 |
| 780.6 | fever nos | 18 | 15.25 |
| 355.9 | mononeuritis of unspecified site | 17 | 14.41 |
| 719.41 | joint pain-shlder | 16 | 13.56 |
| 719.47 | pain in joint, ankle and foot | 16 | 13.56 |
| 780.4 | dizziness and giddiness | 16 | 13.56 |
| 311 | depressive disorder, not elsewhere classified | 15 | 12.71 |
| 477.9 | allergic rhinitis, cause unspecified | 15 | 12.71 |
| 719.46 | joint pain-l/leg | 15 | 12.71 |
| 724.2 | lumbago | 15 | 12.71 |
| 739.1 | somat dysfunc cervic reg | 15 | 12.71 |
| 34 | strep sore throat | 14 | 11.86 |

| | | | |
|---|---|---|---|
| 285.9 | anemia nos | 14 | 11.86 |
| 356.9 | idio periph neurpthy nos | 14 | 11.86 |
| 564 | constipation nos | 14 | 11.86 |
| 719.42 | joint pain-up/arm | 14 | 11.86 |
| 719.43 | joint pain-forearm | 14 | 11.86 |
| 787.02 | nausea alone | 14 | 11.86 |
| 477 | rhinitis due to pollen | 13 | 11.02 |
| 354 | carpal tunnel syndrome | 12 | 10.17 |
| 477.8 | allergic rhinitis nec | 12 | 10.17 |
| 487.1 | flu w resp manifest nec | 12 | 10.17 |
| 724.5 | backache nos | 12 | 10.17 |
| 787.91 | diarrhea | 12 | 10.17 |
| 79.99 | unspecified viral infection | 11 | 9.32 |
| 354.2 | ulnar nerve lesion | 11 | 9.32 |
| 461 | ac maxillary sinusitis | 11 | 9.32 |
| 463 | acute tonsillitis | 11 | 9.32 |
| 473.9 | chronic sinusitis nos | 11 | 9.32 |
| 530.81 | esophageal reflux | 11 | 9.32 |
| 739.2 | somat dysfunc thorac reg | 11 | 9.32 |
| 786.09 | respiratory abnorm nec | 11 | 9.32 |
| 787.01 | nausea with vomiting | 11 | 9.32 |
| 78.1 | viral warts nos | 10 | 8.47 |
| 88.81 | lyme disease | 10 | 8.47 |
| 300.02 | generalized anxiety dis | 10 | 8.47 |
| 314 | attn defic nonhyperact | 10 | 8.47 |
| 382.9 | otitis media nos | 10 | 8.47 |
| 719.44 | pain in joint, hand | 10 | 8.47 |
| 786.05 | shortness of breath | 10 | 8.47 |
| 883 | open wound of finger | 10 | 8.47 |

**Table 3.** Drugs prescribed in 5 years following mononeuritis multiplex diagnosis, observed in at least 10 patients.

| RxNorm ID | Drug name | # Patients | % Patients |
|---|---|---|---|
| 18631 | azithromycin | 41 | 34.75 |
| 723 | amoxicillin | 41 | 34.75 |
| 161 | acetaminophen | 40 | 33.90 |
| 5489 | hydrocodone | 34 | 28.81 |
| 4124 | ethinyl estradiol | 26 | 22.03 |
| 48203 | clavulanate | 23 | 19.49 |
| 8640 | prednisone | 21 | 17.80 |
| 10829 | trimethoprim | 18 | 15.25 |
| 2231 | cephalexin | 18 | 15.25 |

| | | | |
|---|---|---|---|
| 26225 | ondansetron | 18 | 15.25 |
| 41126 | fluticasone | 18 | 15.25 |
| 6387 | lidocaine | 18 | 15.25 |
| 10180 | sulfamethoxazole | 17 | 14.41 |
| 25480 | gabapentin | 17 | 14.41 |
| 2582 | clindamycin | 17 | 14.41 |
| 435 | albuterol | 17 | 14.41 |
| 25037 | cefdinir | 16 | 13.56 |
| 2551 | ciprofloxacin | 16 | 13.56 |
| 5640 | ibuprofen | 16 | 13.56 |
| 748794 | inert ingredients | 16 | 13.56 |
| 6980 | minocycline | 15 | 12.71 |
| 3640 | doxycycline | 14 | 11.86 |
| 10753 | tretinoin | 13 | 11.02 |
| 7514 | norethindrone | 13 | 11.02 |
| 7804 | oxycodone | 13 | 11.02 |
| 8745 | promethazine | 13 | 11.02 |
| 10689 | tramadol | 11 | 9.32 |
| 1418 | benzoyl peroxide | 11 | 9.32 |
| 4450 | fluconazole | 11 | 9.32 |
| 5492 | hydrocortisone | 11 | 9.32 |
| 7258 | naproxen | 11 | 9.32 |
| 7597 | nystatin | 11 | 9.32 |
| 2598 | clonazepam | 10 | 8.47 |
| 2670 | codeine | 10 | 8.47 |
| 6922 | metronidazole | 10 | 8.47 |
| 704 | amitriptyline | 10 | 8.47 |
| 88249 | montelukast | 10 | 8.47 |

### Example consultation report #2

**CLINICAL QUESTION**

Is there a relationship between HSV or VZV infection in patients treated with PD-1/PD-L1 inhibitor immunotherapy?

**HOW WE ASKED THIS QUESTION**

Patients with a history of melanoma are patients with ICD9 code 172 or ICD10 code C43 at any point in their record. Patients treated with a PD-1 or PD-L1 inhibitor are patients whose first prescription record for any drug with RxCUI 1597876, 1547545, 1792776, or 1919503 (with status "inpatient administered", "inpatient prescribed", or "outpatient") occurred after their first melanoma diagnosis, and in 2014 or later. Patients treated with any anti-neoplastic agent (ANA) other than PD-1 or PD-L1 inhibitors are patients whose first prescription record for a drug with ATC class L01 occurred after their first melanoma diagnosis, in 2014 or later, and who were never prescribed a PD-1 or PD-L1 inhibitor.

The outcome of interest is a composite outcome of varicella zoster virus (VZV) or herpes simplex virus (HSV) infection, identified as the first mention of any of ICD9 codes 052, 053, or 054, ICD10 codes B00, B01 or B02, or any lab with LOINC code 16953-2, 16130-7, 16952-4, 16131-5, 16959-9, 16960-7, 11483-5, 21598-8, or 8049-9 and with result "DETECTED".

Note that patients treated with a PD-1 or PD-L1 inhibitor may also have been treated with any other ANA, and that patients who had either VZV or HSV prior to the intervention (PD-1 or PD-L1 inhibitor or any ANA) will be counted as not having experienced the outcome of interest.

Data source: Stanford Hospital & Clinics EHRs from 2014 to 2018.

#### Researcher Interpretation

We identified 587 patients at Stanford who had a history of melanoma and received subsequent anti-neoplastic (ANA) therapy or PD-1/PD-L1 therapy (PD1_INHIBITOR). We defined VZV/HSV reinfection as either a new mention of the associated viral infection ICD codes or PCR positive VZV/HSV in a patient sample anytime after treatment. Matching for patient age, sex, length of the medical record, and year of cohort entry we did not identify any significant difference between VZV/HSV reinfection in the PD1_INHIBITOR cohort.

#### Summary of Cohort

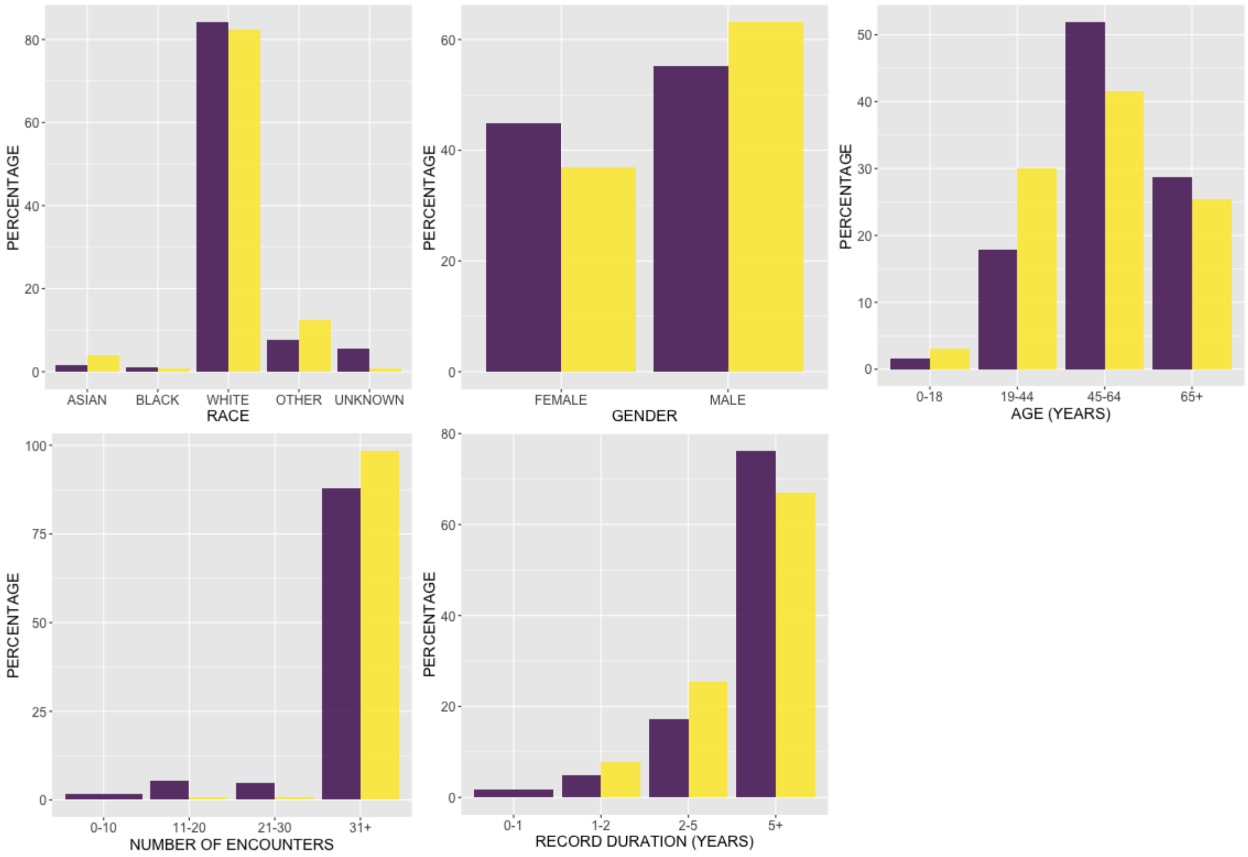


**Figure 1.** Demographics summary of patients with a history of melanoma, treated with any antineoplastic agent other than PD-1 or PD-L1 inhibitors (purple), or with a PD-1 inhibitor (yellow) in 2014 or later.

The following table summarizes the cohort with respect to the various intervention groups and outcomes.

| | **ANA** | **PD1_INHIBITOR** |
|---|---|---|
| **N** | 459 | 128 |
| **Mean age** | 65.2 (13.8) | 61.0 (16.8) |
| **% female** | 44.9 % | 36.7 % |
| **N vzv_hsv (%)** | 12 (2.6%) | 3 (2.3%) |

#### Analysis

We estimated the effect of the treatments relative to the baseline treatment of ANA. We attempt three levels of analysis. First, we perform an unadjusted analysis in which we use the cohorts as is, without any attempt to control for potential confounding. Second, we perform an analysis using a matched cohort in which patients are matched for age, gender, year of entry into the cohort, and record length. Finally, we perform attempt fit a propensity score model using all available data prior to cohort entry in the EHR, and create a matched cohort using the resulting propensity scores. A summary of each cohort is also shown for each comparison and outcome.

The type of analysis performed depends on the type of outcome. For discrete outcomes, we calculate odds ratios and associated confidence intervals. For continuous outcomes, we fit regression models and report the resulting effect estimates and confidence intervals. For time to event or survival outcomes, we show Kaplan-Meier plots along with results from log rank tests for differences in the survival curves. The outcomes and their associated types are shown below.

| OUTCOME | ANALYSIS_TYPE |
|---|---|
| vzv_hsv | discrete |

### 1. Outcome: vzv_hsv

#### 1.1. Outcome: vzv_hsv, Comparison: PD1_INHIBITOR vs ANA

##### *1.1.1. Unadjusted*

| | **ANA** | **PD1_INHIBITOR** |
|---|---|---|
| **N** | 459 | 128 |
| **Mean age (s.d.)** | 65.2 (13.8) | 61.0 (16.8) |
| **% Female** | 44.9 | 36.7 |
| **N w/ outcome** | 12.0 | 3.0 |
| **Mean length of record (s.d.)** | 8.8 (6.5) | 8.0 (6.4) |

| | **Negative** | **Positive** | **Odds.Ratio** | **p.value** |
|---|---|---|---|---|
| **ANA** | 447 | 12 | 1 | NA |
| **PD1_INHIBITOR** | 125 | 3 | 0.894 (0.248, 3.217) | 1 |

##### *1.1.2. Basic Matching*

| | **ANA** | **PD1_INHIBITOR** |
|---|---|---|
| **N** | 128 | 128 |
| **Mean age (s.d.)** | 62.2 (14.5) | 61.0 (16.8) |
| **% Female** | 36.7 | 36.7 |
| **N w/ outcome** | 1.0 | 3.0 |
| **Mean length of record (s.d.)** | 8.1 (6.4) | 8.0 (6.4) |

| | **Negative** | **Positive** | **Odds.Ratio** | **p.value** |
|---|---|---|---|---|
| **ANA** | 127 | 1 | 1 | NA |
| **PD1_INHIBITOR** | 125 | 3 | 3.048 (0.313, 29.698) | 0.622 |

##### *1.1.3. Propensity Score Matching*

Propensity score matching failed for this comparison. This is most often because none of the predictors were useful for predicting the treatment each patient received. This results in a propensity score model in which each patient receives the exact same propensity score.

### Example consultation report #3

**CLINICAL QUESTION**
Between 2013 and 2017, in women, men, adults ≤ 45 years old and adults > 45 years old, how frequently was a positive procalcitonin (defined as a result > 0.5) associated with a positive blood culture?

**HOW WE ASKED THIS QUESTION**
In adult patients (18 years or older), how many procalcitonin lab tests were performed between 2013 and 2017, in women, men, adults ≤ 45 years old and adults > 45 years old? Of those, how many were accompanied by a blood culture within 48 hours (before or after) the procalcitonin lab? How many positive procalcitonin lab test results are accompanied by a positive blood culture, or negative blood culture?

Data source: Stanford Hospital and Clinics electronic health records, 2013-2017.

#### Researcher Interpretation

In women, positive procalcitonin labs are not significantly associated with positive blood cultures (Pearson's Chi-squared test, $X^2$ = 0.03, p = 0.87). In men, positive procalcitonin labs are significantly associated with positive blood cultures (Pearson's Chi-squared test, $X^2$ = 7.86, p < 0.01). In adults ≤ 45 years old, positive procalcitonin labs are not significantly associated with positive blood cultures (Pearson's Chi-squared test, $X^2$ = 1.03, p = 0.31). In adults > 45 years old, positive procalcitonin labs are not significantly associated with positive blood cultures (Pearson's Chi-squared test, $X^2$ = 3.56, p = 0.06).

#### Summary of Cohort

DEFINITIONS

1) Procalcitonin lab: LOINC 33959-8
2) Procalcitonin lab result: >0.5 = POSITIVE; ≤0.5 = NEGATIVE
3) Blood culture: LABS=CULTURE [BLOOD]
4) Blood culture result: NEGATIVE or COAG NEGATIVE STAPHYLOCOCCUS = NEGATIVE; any other result = POSITIVE. If a procalcitonin lab was accompanied by both a negative blood culture result and a positive blood culture result within 48 hours of the procalcitonin lab, this was counted as a positive blood culture.

#### Research walkthrough

POPULATION

There were 6,846 procalcitonin tests performed in women between 2013 and 2017, 9,669 in men, 3067 in adults ≤ 45 years old and 13,302 in adults > 45 years old. There were 13,687 blood cultures performed in women between 2013 and 2017, 15,921 in men, 7,984 in adults ≤ 45 years old and 21,203 in adults > 45 years old.

ANALYSIS

**Women**

Table 1 summarizes the number of positive procalcitonin lab test results accompanied by a positive blood culture within 48 hours, the number of positive procalcitonin lab test results accompanied by a negative blood culture, the number of negative procalcitonin lab test results accompanied by a positive blood culture within 48 hours, and the number of negative procalcitonin lab test results accompanied by a negative blood culture, in women. The majority of procalcitonin labs, with either positive or negative results, were accompanied by a negative blood culture. Positive procalcitonin labs are not significantly associated with positive blood cultures (Pearson's Chi-squared test, $X^2$ = 0.03, p=0.87).

**Table 1.** Association between procalcitonin lab test results and blood cultures taken within 48 hours.

| | | **PROCALCITONIN** | | **TOTAL** |
|---|---|---|---|---|
| | | **POSITIVE** | **NEGATIVE** | |
| **BLOOD CULTURE WITHIN 48 HOURS OF PROCALCITONIN** | **POSITIVE** | 14 | 11 | 25 |
| | **NEGATIVE** | 377 | 344 | 721 |
| **TOTAL** | | 391 | 355 | 746 |

Blood Culture as gold standard:
Sensitivity: 14/25 = 0.56
Specificity: 344/721 = 0.48

Figure 1 summarizes the demographics of patients with a positive procalcitonin lab test result and positive blood culture within 48 hours, negative procalcitonin lab test result and positive blood culture within 48 hours, positive procalcitonin lab test result and negative blood culture within 48 hours, or negative procalcitonin lab test result and negative blood culture within 48 hours.

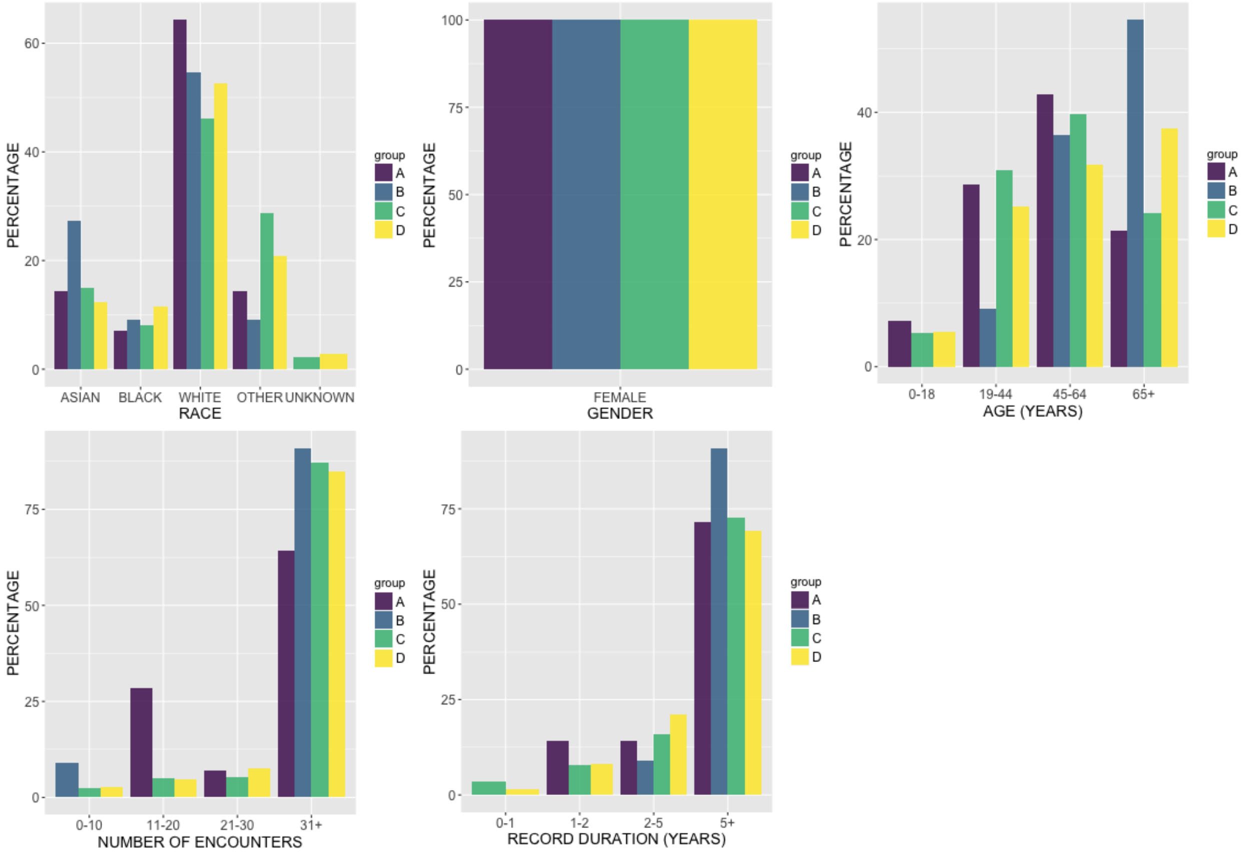


**Figure 1.** Demographics summary of female patients with a positive procalcitonin lab test result and positive blood culture within 48 hours (A), negative procalcitonin lab test result and positive blood culture within 48 hours (B), positive procalcitonin lab test result and negative blood culture within 48 hours (C), or negative procalcitonin lab test result and negative blood culture within 48 hours (D).

**Men**

Table 2 summarizes the number of positive procalcitonin lab test results accompanied by a positive blood culture within 48 hours, the number of positive procalcitonin lab test results accompanied by a negative blood culture, the number of negative procalcitonin lab test results accompanied by a positive blood culture within 48 hours, and the number of negative procalcitonin lab test results accompanied by a negative blood culture, in men. The majority of procalcitonin labs, with either positive or negative results, were accompanied by a negative blood culture. Positive procalcitonin labs are significantly associated with positive blood cultures (Pearson's Chi-squared test, $\boldsymbol{X}^2 = 7.86$, $p < 0.01$).

**Table 2.** Association between procalcitonin lab test results (with a positive result defined as a value ≥ 0.5) and blood cultures taken within 48 hours.

| | | **PROCALCITONIN** | | **TOTAL** |
|---|---|---|---|---|
| | | **POSITIVE** | **NEGATIVE** | |
| **BLOOD CULTURE WITHIN 48 HOURS OF PROCALCITONIN** | **POSITIVE** | 23 | 4 | 27 |
| | **NEGATIVE** | 484 | 378 | 862 |
| **TOTAL** | | 507 | 382 | 889 |

Blood Culture as gold standard:
Sensitivity: 23/27 = 0.85
Specificity: 378/862 = 0.44

Figure 2 summarizes the demographics of patients with a positive procalcitonin lab test result and positive blood culture within 48 hours, negative procalcitonin lab test result and positive blood culture within 48 hours, positive procalcitonin lab test result and negative blood culture within 48 hours, or negative procalcitonin lab test result and negative blood culture within 48 hours.

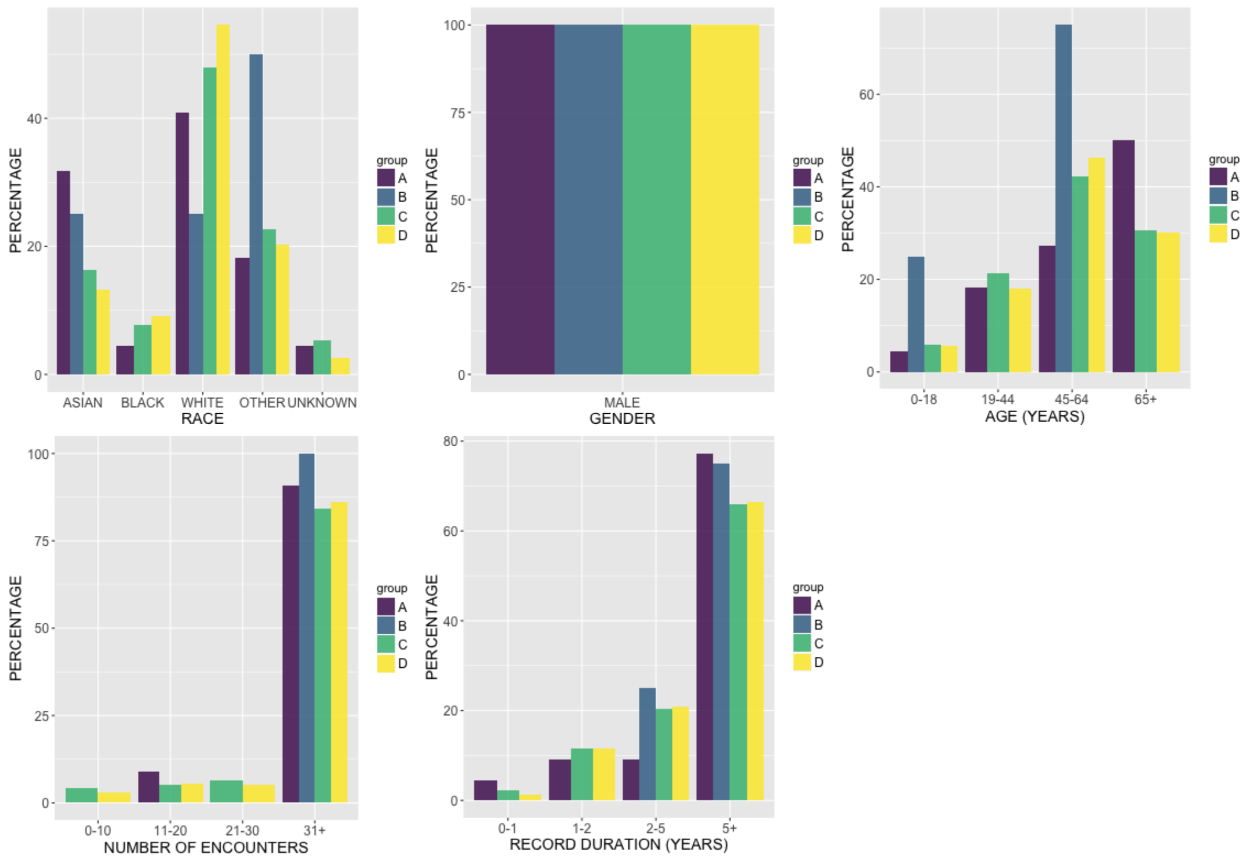


**Figure 2.** Demographics summary of male patients with a positive procalcitonin lab test result and positive blood culture within 48 hours (A), negative procalcitonin lab test result and positive blood culture within 48 hours (B), positive procalcitonin lab test result and negative blood culture within 48 hours (C), or negative procalcitonin lab test result and negative blood culture within 48 hours (D).

**Patients ≤ 45 years of age**

Table 3 summarizes the number of positive procalcitonin lab test results accompanied by a positive blood culture within 48 hours, the number of positive procalcitonin lab test results accompanied by a negative blood culture, the number of negative procalcitonin lab test results accompanied by a positive blood culture within 48 hours, and the number of negative procalcitonin lab test results accompanied by a negative blood culture, in patients between 18 and 45 years of age. The majority of procalcitonin labs, with either positive or negative results, were accompanied by a negative blood culture. Positive procalcitonin labs are not significantly associated with positive blood cultures (Pearson's Chi-squared test, $\boldsymbol{X}^2 = 1.03$, p = 0.31).

**Table 3.** Association between procalcitonin lab test results (with a positive result defined as a value ≥ 2.0) and blood cultures taken within 48 hours.

| | | **PROCALCITONIN** | | **TOTAL** |
|---|---|---|---|---|
| | | **POSITIVE** | **NEGATIVE** | |
| **BLOOD CULTURE WITHIN 48 HOURS OF PROCALCITONIN** | **POSITIVE** | 8 | 2 | 10 |
| | **NEGATIVE** | 199 | 139 | 338 |
| **TOTAL** | | 207 | 141 | 348 |

Blood Culture as gold standard:
Sensitivity: 8/10 = 0.80
Specificity: 139/338 = 0.41

Figure 3 summarizes the demographics of patients between 18 and 45 years of age with a positive procalcitonin lab test result and positive blood culture within 48 hours, negative procalcitonin lab test result and positive blood culture within 48 hours, positive procalcitonin lab test result and negative blood culture within 48 hours, or negative procalcitonin lab test result and negative blood culture within 48 hours.

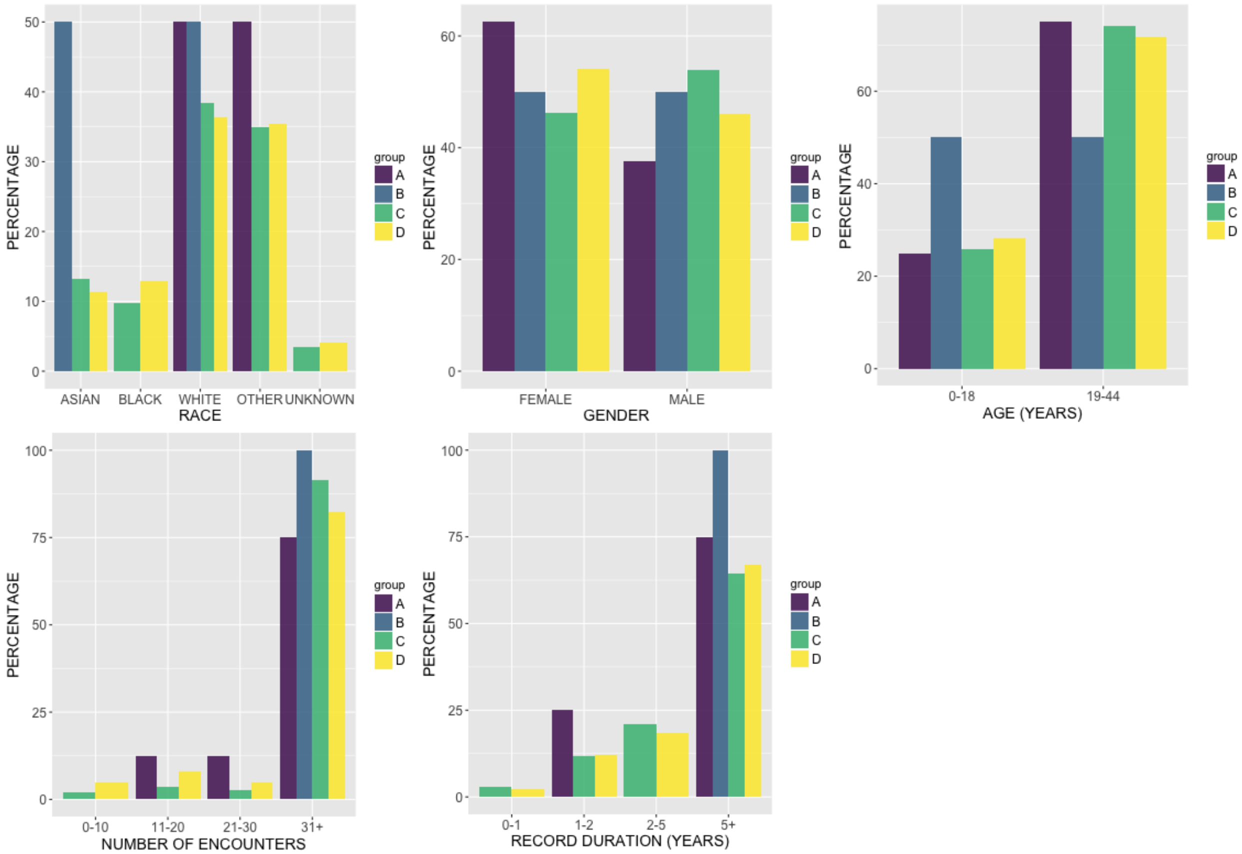


**Figure 3.** Demographics summary of patients aged 18 - 45 years with a positive procalcitonin lab test result and positive blood culture within 48 hours (A), negative procalcitonin lab test result and positive blood culture within 48 hours (B), positive procalcitonin lab test result and negative blood culture within 48 hours (C), or negative procalcitonin lab test result and negative blood culture within 48 hours (D).

**<u>Patients > 45 years of age</u>**

Table 4 summarizes the number of positive procalcitonin lab test results accompanied by a positive blood culture within 48 hours, the number of positive procalcitonin lab test results accompanied by a negative blood culture, the number of negative procalcitonin lab test results accompanied by a positive blood culture within 48 hours, and the number of negative procalcitonin lab test results accompanied by a negative blood culture, in patients over 45 years of age. The majority of procalcitonin labs, with either positive or negative results, were accompanied by a negative blood culture. Positive procalcitonin labs are not significantly associated with positive blood cultures (Pearson's Chi-squared test, $\boldsymbol{X}^2 = 3.56$, p = 0.06).

**Table 4.** Association between procalcitonin lab test results (with a positive result defined as a value ≥ 2.0) and blood cultures taken within 48 hours.

| | | PROCALCITONIN | | TOTAL |
|---|---|---|---|---|
| | | POSITIVE | NEGATIVE | |
| BLOOD CULTURE WITHIN 48 HOURS OF PROCALCITONIN | POSITIVE | 29 | 13 | 42 |
| | NEGATIVE | 651 | 576 | 1227 |
| TOTAL | | 680 | 589 | 1269 |

Blood Culture as gold standard:
Sensitivity: 29/42 = 0.69
Specificity: 576/1227 = 0.47

Figure 4 summarizes the demographics of patients older than 45 years with a positive procalcitonin lab test result and positive blood culture within 48 hours, negative procalcitonin lab test result and positive blood culture within 48 hours, positive procalcitonin lab test result and negative blood culture within 48 hours, or negative procalcitonin lab test result and negative blood culture within 48 hours.

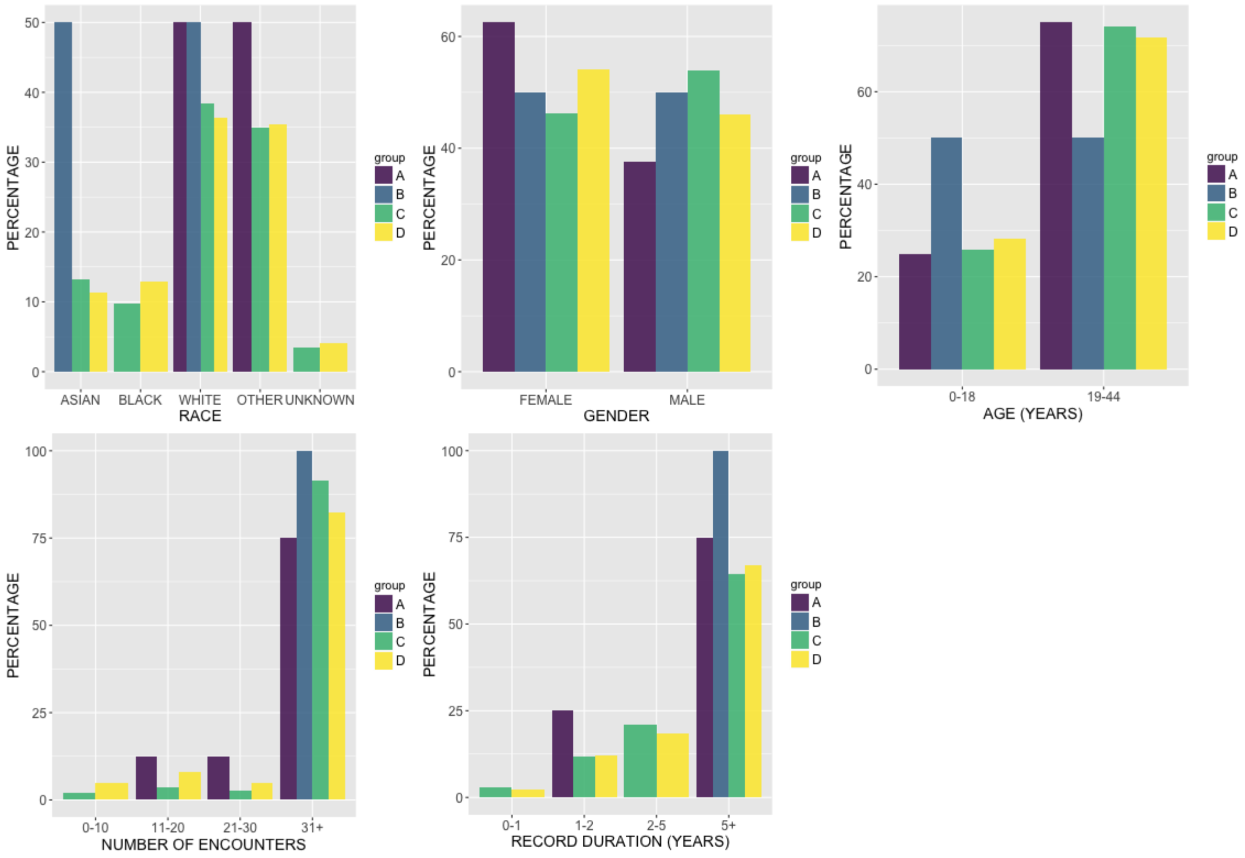


**Figure 4.** Demographics summary of patients older than 45 years with a positive procalcitonin lab test result and positive blood culture within 48 hours (A), negative procalcitonin lab test result and positive blood culture within 48 hours (B), positive procalcitonin lab test result and negative blood culture within 48 hours (C), or negative procalcitonin lab test result and negative blood culture within 48 hours (D).

Table 5 summarizes the number of procalcitonin and blood culture lab test records with varying filters applied, to illustrate how the results in Tables 1-4 were obtained. For example, the final number of procalcitonin lab test results in women included in the analysis was 746, after filtering to procalcitonin labs accompanied by at least one blood culture result within 48 hours (before or after) and where the patient was less than 90 years of age. The age filter was applied because once a patient reaches the age of 90, all records after that point are stated to occur at age 90 (per HIPAA requirements), so it is not possible to accurately identify blood cultures that occur within 48 hours.

**Table 5.** The number of records for procalcitonin and blood culture lab tests, with increasing filters applied, and with different result flags. ANY = a positive or negative test result; POSITIVE/POS = positive test result; NEGATIVE/NEG = negative test result.

| LAB TEST | FILTER | RESULT | COUNT IN COHORT | | | |
|---|---|---|---|---|---|---|
| | | | **Women** | **Men** | **≤45** | **>45** |
| PROCALCITONIN | NONE | ANY | 6846 | 9669 | 3067 | 13302 |
| PROCALCITONIN | HAS VALID RESULT VALUE | ANY | 6434 | 9207 | 3007 | 12864 |
| PROCALCITONIN | HAS VALID RESULT VALUE | POSITIVE | **3666** | **5514** | **1857** | **7374** |
| PROCALCITONIN | HAS VALID RESULT VALUE | NEGATIVE | **2768** | **3693** | **1150** | **5490** |
| PROCALCITONIN | HAS VALID RESULT VALUE; BLOOD CULTURE DONE WITHIN 48 HOURS | ANY | 788 | 926 | 348 | 1348 |
| PROCALCITONIN | HAS VALID RESULT VALUE; BLOOD CULTURE DONE WITHIN 48 HOURS; AGE < 90 YEARS | ANY | 746 | 889 | 348 | 1269 |
| BLOOD CULTURE | HAS VALID RESULT VALUE | ANY | 13687 | 15921 | 7984 | 21203 |
| BLOOD CULTURE | HAS VALID RESULT VALUE | POSITIVE | 386 | 444 | 172 | 645 |
| BLOOD CULTURE | HAS VALID RESULT VALUE | NEGATIVE | 13301 | 15477 | 7812 | 20558 |
| BLOOD CULTURE | HAS VALID RESULT VALUE; DONE WITHIN 48 HOURS OF PROCALCITONIN | ANY | 961 | 1162 | 436 | 1660 |
| BLOOD CULTURE | HAS VALID RESULT VALUE; DONE WITHIN 48 HOURS OF PROCALCITONIN; AGE < 90 YEARS | ANY | 916 | 1114 | 436 | 1567 |
| PROCALCITONIN + BLOOD CULTURE | HAS VALID RESULT VALUE; DONE WITHIN 48 HOURS OF EACH OTHER; AGE < 90 YEARS | POS/POS | **14** | **23** | **8** | **29** |
| PROCALCITONIN + BLOOD CULTURE | HAS VALID RESULT VALUE; DONE WITHIN 48 HOURS OF EACH OTHER; AGE < 90 YEARS | POS/NEG | **377** | **484** | **199** | **651** |
| PROCALCITONIN + BLOOD CULTURE | HAS VALID RESULT VALUE; DONE WITHIN 48 HOURS OF EACH OTHER; AGE < 90 YEARS | NEG/POS | **11** | **4** | **2** | **13** |
| PROCALCITONIN + BLOOD CULTURE | HAS VALID RESULT VALUE; DONE WITHIN 48 HOURS OF EACH OTHER; AGE < 90 YEARS | NEG/NEG | **344** | **378** | **139** | **576** |
